# Risk factors for hospitalisation and death due to COVID-19 during endemic Omicron circulation, a population-based cohort study

**DOI:** 10.1101/2025.05.05.25326987

**Authors:** Ulrika Marking, Erik Wahlström, Johanna Holm, Sten Walther, Håkan Hanberger, Kristoffer Strålin

## Abstract

**Background:** Knowledge about risk factors for hospitalisation and death due to COVID-19 during the Omicron period, beyond the initial Omicron wave, remains limited. Therefore, to support vaccination strategies and antiviral treatment recommendations, we aimed to investigate the significance of various risk factors and their association with hospitalisation and death from COVID-19 during the Omicron period.

**Method:** Nationwide registry data on the Swedish adult population was compiled for the period June 6, 2022, to March 31, 2023. Frequency of hospitalisation with a primary discharge diagnosis of COVID-19 and death with COVID-19 specified as the underlying cause of death on the death certificate were analysed. Individual data were stratified by age, vaccination status, Charlson Comorbidity Index, and specific comorbidities. Poisson regression was used to estimate incidence risk ratios of independent risk factors for hospitalisation and death including age, sex, comorbidities, care dependency and socioeconomic factors.

**Results:** Among the more than 8,3 million individuals in the study cohort, 13 941 hospitalisations and 2 309 deaths due to COVID-19 were recorded during the study period. Multivariable analysis revealed that high age was the strongest risk factors for both hospitalisation and death. Advanced comorbidity was the second strongest risk factor for hospitalisation, and care dependency was the second strongest risk factor for death. Among comorbidities, recent chemotherapy, Immune Deficiency, Multiple Sclerosis, and Chronic Kidney Disease stage 5 were the strongest risk factors for hospitalisation and death.

**Conclusion:** Also in the current Omicron endemicity, advanced age remains the strongest risk factor for hospitalisation and death due to COVID-19, accompanied by advanced comorbidity and care dependency. These findings can guide recommendations for vaccination strategies and antiviral treatment, helping to protect those most at risk.

## Introduction

Ancestral SARS-CoV-2 were, just like the initially dominant variants of concern alpha and delta more prone to cause severe disease and death than the later emerging Omicron variant (1–4). Omicron appearance coincided with a high vaccine uptake in the population, but also among unvaccinated the Omicron variant exhibited considerably less virulence (2–5). Several explanations have been brought forward, such as a tropism more restricted to the upper respiratory tract than the lungs (6) and reduced fusogenicity of the spike protein leading to less extensive tissue damage (7). Regardless of mechanism, the rapid omicron transmission led to wide-spread infection acquired immunity, significantly shifting the risk landscape attributed to SARS-CoV-2.

With increased hybrid immunity from a combination of vaccination and one or multiple infections the risk of severe COVID-19 outcomes is reduced considerable, and interventions to mitigate disease burden may need adaptations. Detailed knowledge of patient factors increasing the risk for severe COVID-19 is essential to inform public health decisions and mitigate risks of infection among the relevant vulnerable groups. This study aims to assess risk factors for hospitalisation and death due to COVID-19 in a setting reflecting the endemic phase of SARS-CoV-2 Omicron circulation in the Swedish population, with study period starting in June 2022, after 6 months of Omicron dominance.

## Methods

### Study population and design

This observational study included all adult (≥ 18 years) individuals registered as residents in Sweden as of December 31, 2021, according to the Total Population Register, Statistics Sweden. Baseline characteristics were described, and the population was followed from June 10^th^ 2022 to March 31^st^ 2023, covering a period of almost 10 months. Outcome data, including hospitalisation for COVID-19 and death due to COVID-19, were registered.

Nationwide registry data were compiled by the Swedish National Board of Health and Welfare using the unique national personal identification number as previously described (5). Briefly, data on comorbidity, care dependency, hospitalisations, mortality, and cause of death were obtained from health registers maintained by the National Board of Health and Welfare, data on SARS-CoV-2 vaccination were obtained from registers held by Public Health Agency of Sweden as previously described, and data on level of education and socio-economic index of residential area was obtained from Statistics Sweden (5).

### Covariate data

Major comorbidities were identified and defined using 1) diagnoses recorded at discharge from hospitalisation or specialised out-patient care; and/or 2) prescribed drugs for chronic conditions commonly managed in primary care dispensed during the year preceding study start; and/or 3) the National Cancer register (Supplementary Table S1). A 30-day wash-out window prior to index admission date was applied. Codes of the 10^th^ International Statistical Classification of Diseases (ICD-10) and Anatomic Therapeutic Chemical (ATC) classifications, and health care procedures defining each comorbidity are provided in Supplementary Table S1. Diagnoses were sourced from the National Patient Register (held by the National Board of Health and Welfare) which contains comprehensive data on diagnoses recorded at hospital discharge or specialized out-patient departments. Data on drugs dispensed were obtained from the Swedish Prescribed Drug Register (held by the National Board of Health and Welfare), which contains all prescribed and pharmacy-dispensed medicines in the community classified according to the ATC Classification System. The register has almost complete coverage (data missing < 1%).

A Charlson Comorbidity Index (CCI) was calculated for all individuals based on the comorbidities registered in the National Patient Register (8) in five years prior to index date. In addition, to study specific comorbidities as exposures in relation to the combined comorbidities of patients in the present study, we calculated a modified CCI, in which all comorbidities of the patients except from the specific exposure of interest were included.

Vaccination data was obtained from the National Vaccination Register, maintained by the Public Health Agency of Sweden, which has mandatory reporting on COVID-19 vaccination. The majority (88.5%) of the Swedish adult population had received at least one dose vaccine against COVID-19 by June 2022 (9).

Socioeconomic index of area of residency as determined yearly by Statistics Sweden was employed to capture socioeconomic status. The measure is based on 1) proportion of citizens with completed primary school; 2) proportion of citizens with a low economic standard based on household income; and 3) proportion of citizens either unemployed for >6 months or with welfare benefits. Education level was obtained from the Education Register held by Statistics Sweden.

Information on care dependency (long-term care facilities for elderly (LTCF), home-based care, or supportive housing for persons with functional impairments) prior to index date was obtained from the Care and Social Services for the Elderly and for Persons with Impairments Register and the National Register of Municipal Support and Service for Persons with Certain Functional Impairment, held by the National Board of Health and Welfare.

### Outcomes

Outcome measures were hospitalisation for COVID-19 defined as hospitalisation with a COVID-19 diagnosis as the primary diagnosis at discharge; and death from COVID-19, defined as death with COVID-19 specified as the underlying cause of death on the death certificate. Both outcomes were binary.

### Statistical analysis

Poisson regression was used to estimate Incidence rate ratios (IRR) for a range of factors associated to one or both outcomes (hospitalisation and death due to COVID-19) using different models. Separate models were created for each exposure – outcome combination. We estimated the effect of each comorbidity in models adjusted for *age* (in 5-year intervals), *sex* (male/female), time since most recent *SARS-CoV-2 vaccination* (<6 months, 6-12 months, 13-24 months, unvaccinated (including vaccination more than 24 months prior to start of study period)), modified *CCI* (0, 1-2, 3-5, 6+), *calendar time* (in 7-day intervals), *socioeconomic index* of area of residency (5 categories), *education* (primary school, secondary school, university/college < 3yrs, university/college > 3 yrs) and *care dependency*, all modelled as main effects. To assess the IRR associated to risk factors included in all models, specific models were created without a specific comorbidity variable but with the unmodified CCI for both study outcomes. Crude analysis of the outcomes (hospitalisation for COVID-19 and death due to COVID-19) are reported as Incidence rate per 1000 person-years.

Registry data were complete on all variables except for education (2.1%), and complete case analysis was applied in all regression models. Sensitivity analyses were performed comparing adjusted IRR when including cases without education data and treating them as an additional category.

All data management and statistical analyses was performed using SAS Software SAS Enterprise Guide version 8.3 (SAS institute, Cary, NC, USA).

### Ethical permission

The study was approved by the Swedish Ethics Review Authority, Uppsala, Sweden (Dnr 2020-04278).

## Results

Among more than 8,3 million individuals in the study cohort, 13 941 hospitalisations and 2 309 deaths due to COVID-19 were recorded during the study period. Table 1 shows characteristics of individuals with or without hospitalisation for COVID-19 and with or without death due to COVID-19 during the study period. As noted, both hospitalisation and death were more frequent among males than females and among individuals ≥ 80 years of age compared with younger individuals.

**Table 1.** Distribution of covariates and sociodemographic characteristics of study cohort per 1000 person-years.

|  | Total |  | Hospitalised due to COVID-19 during study period |  |  |  | Death due to COVID-19 during study period |  |  |  |
| --- | --- | --- | --- | --- | --- | --- | --- | --- | --- | --- |
|  |  |  | No |  | Yes |  | No |  | Yes |  |
|  | N | % | N | % | N | % | N | % | N | % |
| **Total** | 8 328 797 | 100 | 8 314 856 | 100 | 13 941 | 100 | 8 326 488 | 100 | 2 309 | 100 |
| **Sex** |  |  |  |  |  |  |  |  |  |  |
| Male | 4 168 606 | 50,1 | 4 160 698 | 50,0 | 7 908 | 56,7 | 4 167 262 | 50,0 | 1 344 | 58,2 |
| Female | 4 160 191 | 49,9 | 4 154 158 | 50,0 | 6 033 | 43,3 | 4 159 226 | 50,0 | 965 | 41,8 |
| **Age** |  |  |  |  |  |  |  |  |  |  |
| 18-64 | 6 136 278 | 73,7 | 6 134 425 | 73,8 | 1 853 | 13,3 | 6 136 205 | 73,7 | 73 | 3,2 |
| 65-79 | 1 579 071 | 19,0 | 1 574 128 | 18,9 | 4 943 | 35,5 | 1 578 522 | 19,0 | 549 | 23,8 |
| 80+ | 613 448 | 7,4 | 606 303 | 7,3 | 7 145 | 51,3 | 611 761 | 7,3 | 1 687 | 73,1 |
| 18-29 | 1 467 573 | 17,6 | 1 467 385 | 17,6 | 188 | 1,3 | 1 467 572 | 17,6 | 1 | 0,0 |
| 30-39 | 1 459 280 | 17,5 | 1 459 038 | 17,5 | 242 | 1,7 | 1 459 273 | 17,5 | 7 | 0,3 |
| 40-49 | 1 294 586 | 15,5 | 1 294 307 | 15,6 | 279 | 2,0 | 1 294 579 | 15,5 | 7 | 0,3 |
| 50-59 | 1 338 491 | 16,1 | 1 337 867 | 16,1 | 624 | 4,5 | 1 338 464 | 16,1 | 27 | 1,2 |
| 60-69 | 1 121 651 | 13,5 | 1 120 296 | 13,5 | 1 355 | 9,7 | 1 121 551 | 13,5 | 100 | 4,3 |
| 70-79 | 1 033 768 | 12,4 | 1 029 660 | 12,4 | 4 108 | 29,5 | 1 033 288 | 12,4 | 480 | 20,8 |
| 80-89 | 498 194 | 6,0 | 492 937 | 5,9 | 5 257 | 37,7 | 497 233 | 6,0 | 961 | 41,6 |
| 90+ | 115 254 | 1,4 | 113 366 | 1,4 | 1 888 | 13,5 | 114 528 | 1,4 | 726 | 31,4 |
| **Care Dependency** |  |  |  |  |  |  |  |  |  |  |
| Self-housing, no care | 8 039 609 | 96,5 | 8 030 434 | 96,6 | 9 175 | 65,8 | 8 038 830 | 96,5 | 779 | 33,7 |
| Home based care | 173 958 | 2,1 | 170 111 | 2,0 | 3 847 | 27,6 | 173 247 | 2,1 | 711 | 30,8 |
| Supportive housing for persons with functional impairments | 29 357 | 0,4 | 29 254 | 0,4 | 103 | 0,7 | 29 343 | 0,4 | 14 | 0,6 |
| Long-term care facilities for elderly | 85 873 | 1,0 | 85 057 | 1,0 | 816 | 5,9 | 85 068 | 1,0 | 805 | 34,9 |
| **Comorbidities (Charlson Comorbidity Index)** |  |  |  |  |  |  |  |  |  |  |
| CCI 0 | 7 111 699 | 85,4 | 7 107 080 | 85,5 | 4 619 | 33,1 | 7 111 043 | 85,4 | 656 | 28,4 |
| CCI 1-2 | 937 382 | 11,3 | 932 393 | 11,2 | 4 989 | 35,8 | 936 485 | 11,2 | 897 | 38,8 |
| CCI 3-5 | 195 581 | 2,3 | 192 389 | 2,3 | 3 192 | 22,9 | 195 015 | 2,3 | 566 | 24,5 |
| CCI 6+ | 84 135 | 1,0 | 82 994 | 1,0 | 1 141 | 8,2 | 83 945 | 1,0 | 190 | 8,2 |
| **Diabetes** |  |  |  |  |  |  |  |  |  |  |
| No diabetes | 7 714 076 | 92,6 | 7 703 876 | 92,7 | 10 200 | 73,2 | 7 712 389 | 92,6 | 1 687 | 73,1 |
| Diabetes without complications | 480 945 | 5,8 | 478 574 | 5,8 | 2 371 | 17,0 | 480 543 | 5,8 | 402 | 17,4 |
| Diabetes with complications | 133 776 | 1,6 | 132 406 | 1,6 | 1 370 | 9,8 | 133 556 | 1,6 | 220 | 9,5 |
| **Hypertension** |  |  |  |  |  |  |  |  |  |  |
| No hypertension | 6 217 201 | 74,6 | 6 213 944 | 74,7 | 3 257 | 23,4 | 6 216 714 | 74,7 | 487 | 21,1 |
| Medication only | 1 324 848 | 15,9 | 1 321 114 | 15,9 | 3 734 | 26,8 | 1 324 195 | 15,9 | 653 | 28,3 |
| Essential hypertension | 781 146 | 9,4 | 774 240 | 9,3 | 6 906 | 49,5 | 779 986 | 9,4 | 1 160 | 50,2 |
| Hypertension with complications or secondary hypertension | 5 602 | 0,1 | 5 558 | 0,1 | 44 | 0,3 | 5 593 | 0,1 | 9 | 0,4 |
| **Pulmonary Disease** |  |  |  |  |  |  |  |  |  |  |
| No pulmonary disease | 7 597 353 | 91,2 | 7 586 829 | 91,2 | 10 524 | 75,5 | 7 595 553 | 91,2 | 1 800 | 78,0 |
| Mild pulmonary disease | 638 528 | 7,7 | 636 829 | 7,7 | 1 699 | 12,2 | 638 284 | 7,7 | 244 | 10,6 |
| Severe pulmonary disease | 92 916 | 1,1 | 91 198 | 1,1 | 1 718 | 12,3 | 92 651 | 1,1 | 265 | 11,5 |
| **Cronic Kidney Disease** |  |  |  |  |  |  |  |  |  |  |
| Not CKD 3-5 | 8 282 450 | 99,4 | 8 269 474 | 99,5 | 12 976 | 93,1 | 8 280 301 | 99,4 | 2 149 | 93,1 |
| CKD 3 | 20 171 | 0,2 | 19 845 | 0,2 | 326 | 2,3 | 20 120 | 0,2 | 51 | 2,2 |
| CKD 4 | 15 596 | 0,2 | 15 260 | 0,2 | 336 | 2,4 | 15 533 | 0,2 | 63 | 2,7 |
| CKD 5 | 10 580 | 0,1 | 10 277 | 0,1 | 303 | 2,2 | 10 534 | 0,1 | 46 | 2,0 |
| **Other Comorbidities** |  |  |  |  |  |  |  |  |  |  |
| Ischemic Heart Disease | 317 766 | 3,8 | 314 525 | 3,8 | 3 241 | 23,2 | 317 200 | 3,8 | 566 | 24,5 |
| Heart Failure | 143 908 | 1,7 | 141 395 | 1,7 | 2 513 | 18,0 | 143 402 | 1,7 | 506 | 21,9 |
| Chronic Liver Disease | 24 540 | 0,3 | 24 372 | 0,3 | 168 | 1,2 | 24 516 | 0,3 | 24 | 1,0 |
| Multiple Sklerosis | 20 951 | 0,3 | 20 780 | 0,2 | 171 | 1,2 | 20 939 | 0,3 | 12 | 0,5 |
| Obesity | 177 556 | 2,1 | 176 969 | 2,1 | 587 | 4,2 | 177 499 | 2,1 | 57 | 2,5 |
| Psychiatric Disorder | 95 726 | 1,1 | 95 381 | 1,1 | 345 | 2,5 | 95 669 | 1,1 | 57 | 2,5 |
| Rheumatoid Arthritis | 66 579 | 0,8 | 66 027 | 0,8 | 552 | 4,0 | 66 484 | 0,8 | 95 | 4,1 |
| Immune Deficiency | 53 620 | 0,6 | 52 618 | 0,6 | 1 002 | 7,2 | 53 478 | 0,6 | 142 | 6,1 |
| Chemotherapy (at any point during study period) | 26 065 | 0,3 | 25 713 | 0,3 | 352 | 2,5 | 26 027 | 0,3 | 38 | 1,6 |
| **Education** |  |  |  |  |  |  |  |  |  |  |
| Primary | 1 449 102 | 17,4 | 1 444 123 | 17,4 | 4 979 | 35,7 | 1 448 123 | 17,4 | 979 | 42,4 |
| Secondary | 3 527 936 | 42,4 | 3 522 197 | 42,4 | 5 739 | 41,2 | 3 527 077 | 42,4 | 859 | 37,2 |
| University less than 3 years | 1 204 519 | 14,5 | 1 203 260 | 14,5 | 1 259 | 9,0 | 1 204 336 | 14,5 | 183 | 7,9 |
| University 3 years or more | 1 971 358 | 23,7 | 1 969 669 | 23,7 | 1 689 | 12,1 | 1 971 119 | 23,7 | 239 | 10,4 |
| Data missing | 175 882 | 2,1 | 175 607 | 2,1 | 275 | 2,0 | 175 833 | 2,1 | 49 | 2,1 |
| **Socioeconomic index of area of** |  |  |  |  |  |  |  |  |  |  |
| Significant challenges | 414 230 | 5,0 | 413 549 | 5,0 | 681 | 4,9 | 414 131 | 5,0 | 99 | 4,3 |
| Some challenges | 683 718 | 8,2 | 682 422 | 8,2 | 1 296 | 9,3 | 683 481 | 8,2 | 237 | 10,3 |
| Mixed area | 1 893 178 | 22,7 | 1 889 386 | 22,7 | 3 792 | 27,2 | 1 892 513 | 22,7 | 665 | 28,8 |
| Favourable conditions | 4 546 201 | 54,6 | 4 539 026 | 54,6 | 7 175 | 51,5 | 4 545 050 | 54,6 | 1 151 | 49,8 |
| Very favourable conditions | 791 470 | 9,5 | 790 473 | 9,5 | 997 | 7,2 | 791 313 | 9,5 | 157 | 6,8 |

Time since most recent vaccination varied over the study period and is presented in Figure 1A. Rates of hospitalisation due to COVID-19 during the study period are presented in Figure 1B.

**Figure 1.**
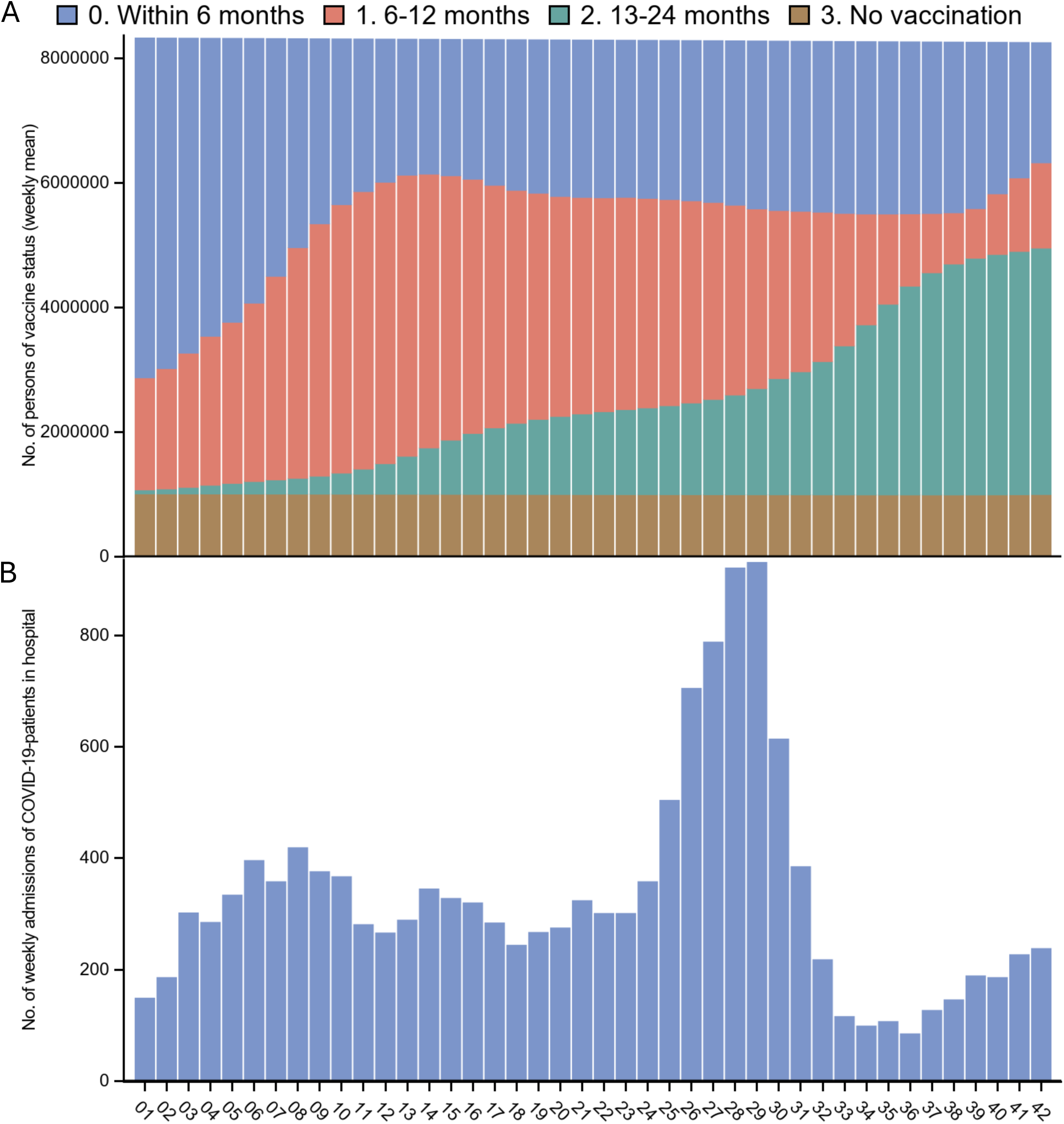
(A) Time since most recent vaccination in study cohort and (B) number of patients hospitalized during study period as a surrogate of SARS-CoV-2 transmission.

In multivariable regression analyses, age remained a significant risk factor, with individuals aged 80–84 exhibiting a 7.1-fold increased risk of hospitalisation (95% CI: 6.5–7.9) (Figure 2A) and a 15.6-fold higher risk of death (95% CI: 10.8–22.6) compared to those aged 60–64 (Figure 3A). A high Charlson Comorbidity Index (CCI) of 3–5 compared to 0, was also a strong predictor of hospitalisation (IRR 4.9 [95% CI 4.7–5.2]), though its association with death was comparatively weaker (IRR 2.7 [95% CI 2.5–3.1]). However, care dependency was strongly associated with death (IRR 12.2 [95% CI 10.8–13.7] for nursing home and IRR 9.5 [95% CI 5.1–17.8] for supportive housing).

**Figure 2.**
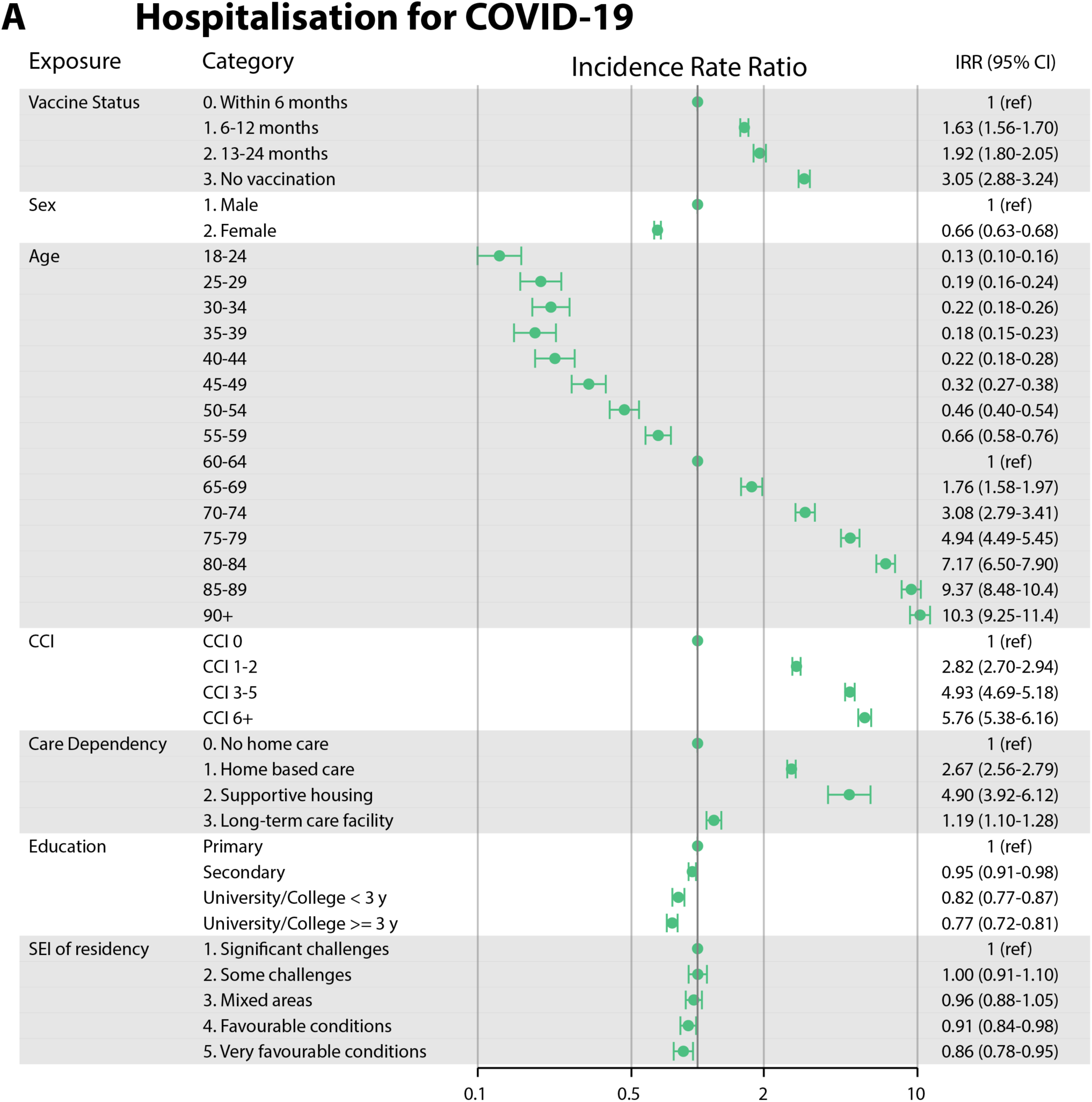
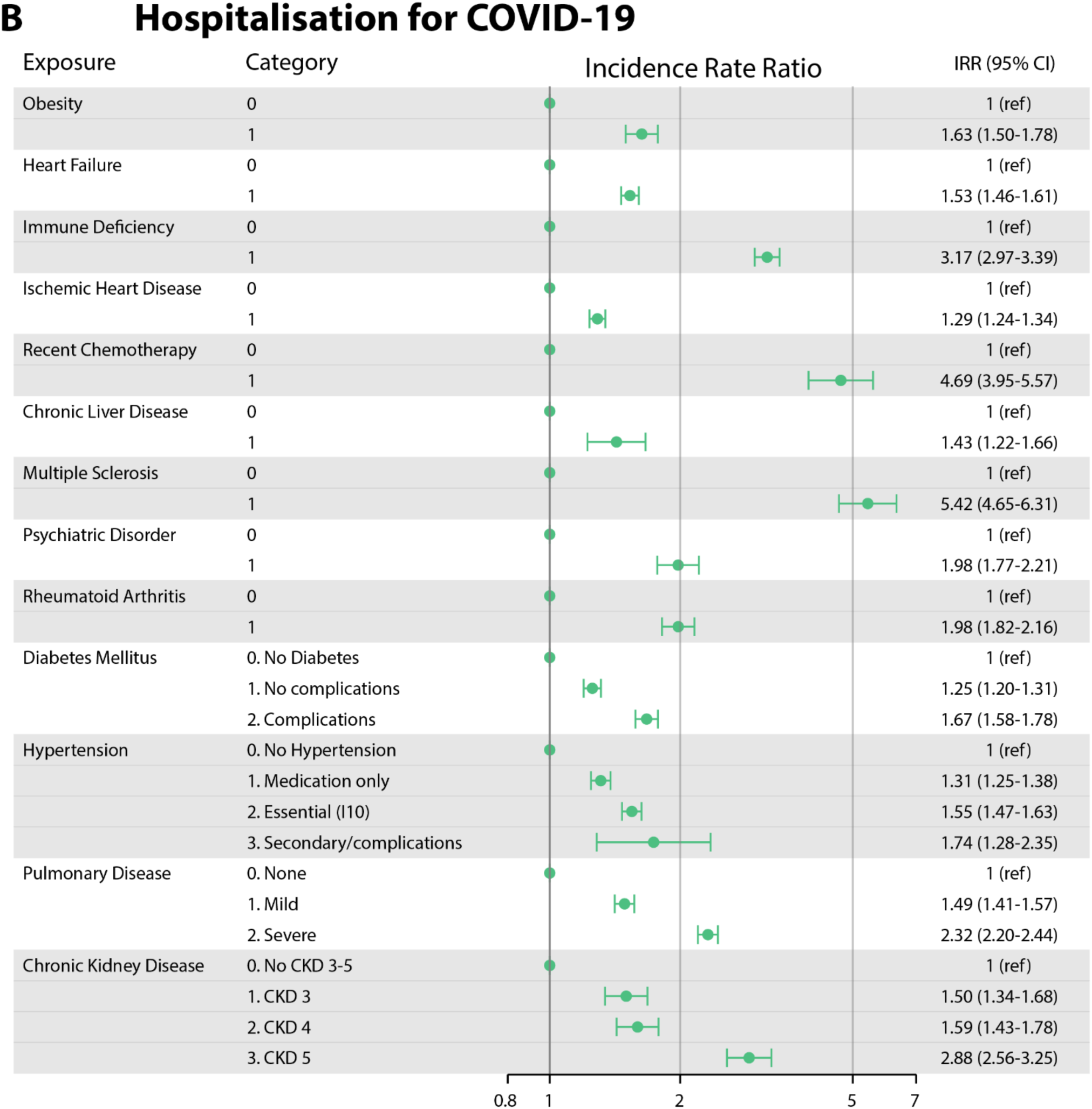
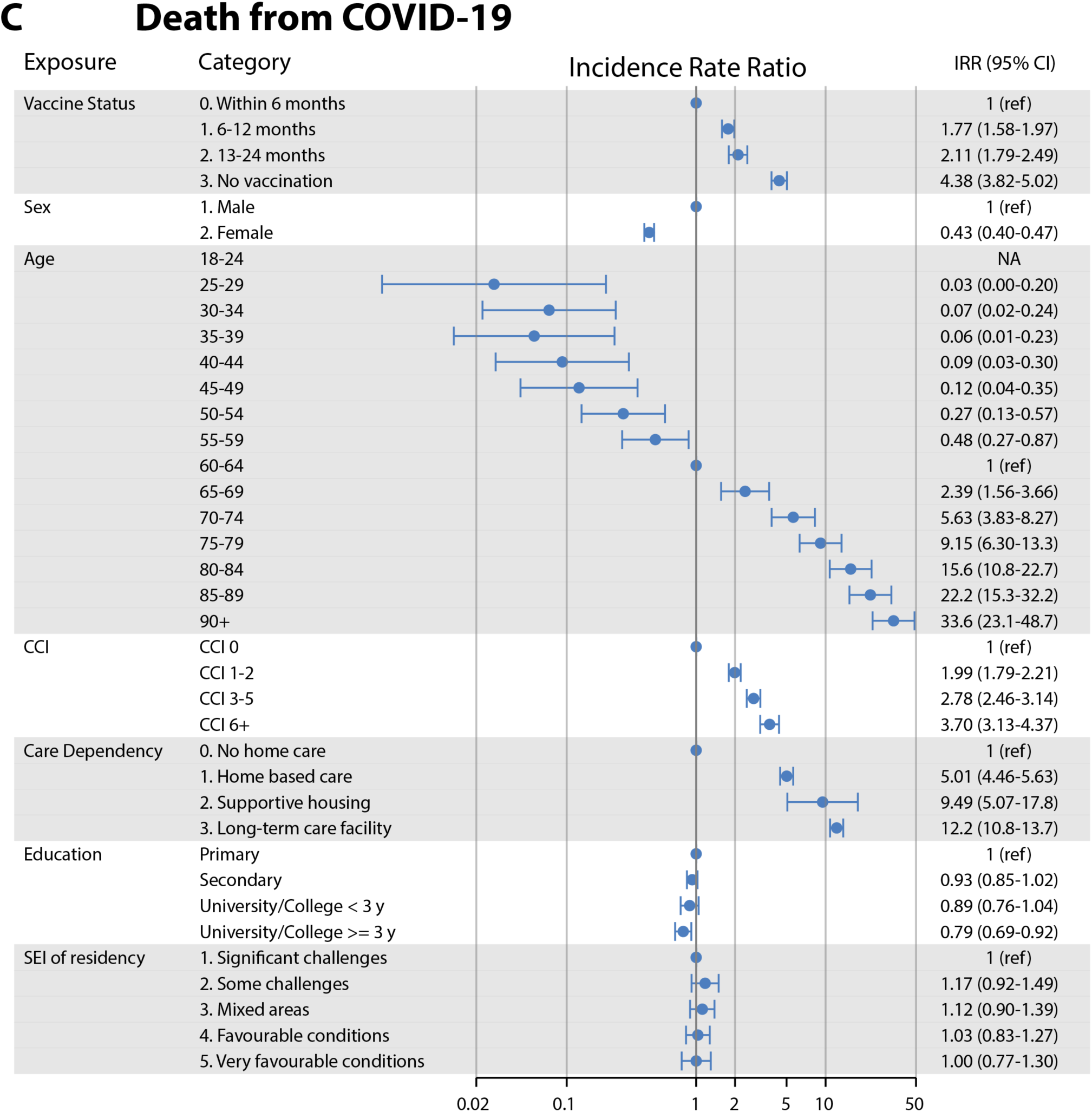
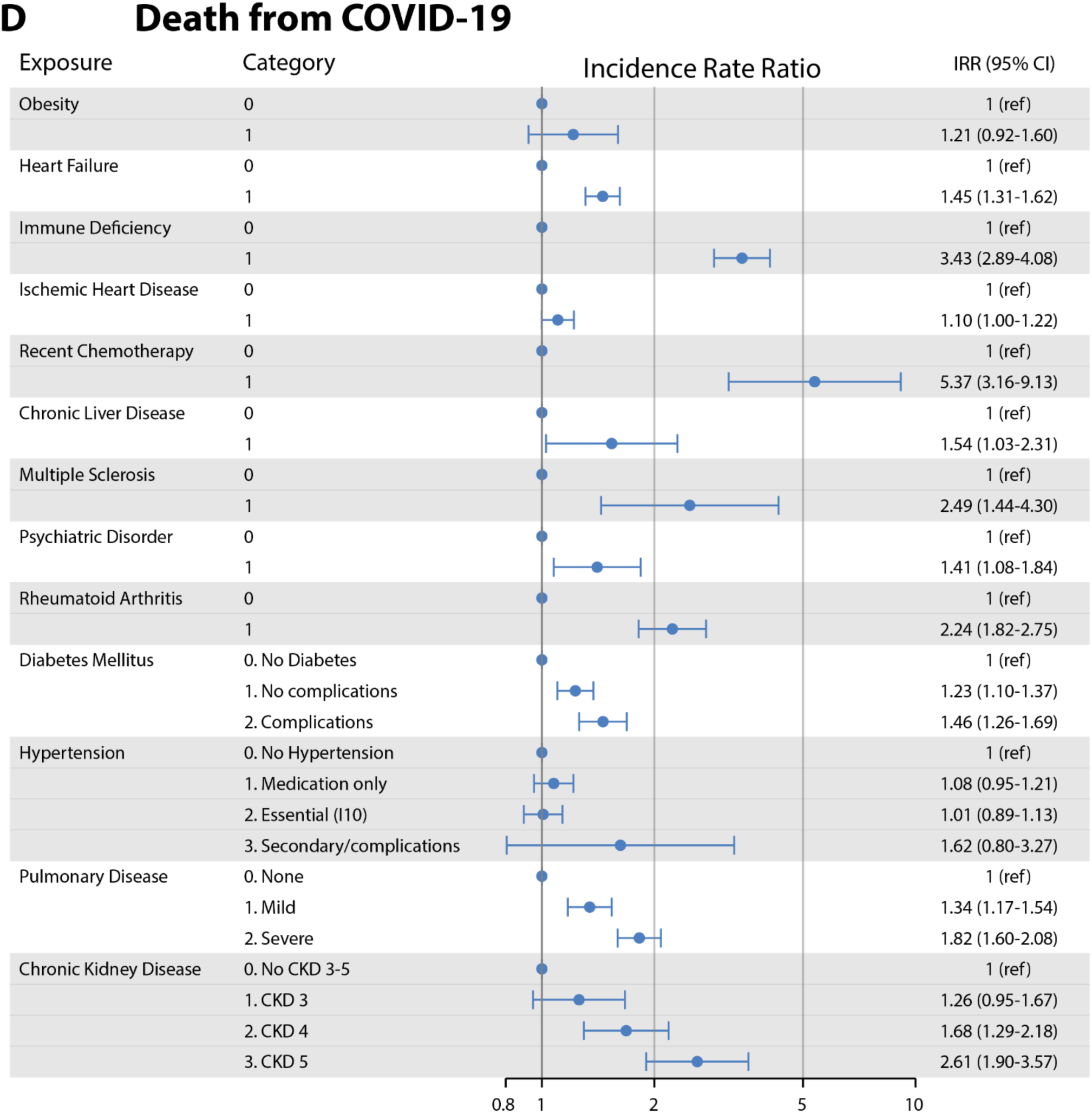
Forest plots of incidence rate ratios with 95% confidence intervals, estimated from multivariable regression models for hospitalisation (A, B) and death (C, D) due to COVID-19. The overall models (A, C) include vaccine status, sex, age, Charlson Comorbidity Index (CCI), care dependency, education, and socioeconomic index (SEI). The disease specific models (B, D) include adjustment for age, sex, vaccination status, modified CCI (total CCI apart from the score rendered by the specific disease), calendar time, care dependency, education, and socioeconomic index, all modelled as main effects. CCI; Charlson Comorbidity Index, SEI; socioeconomic Index

Among specific diseases, recent chemotherapy, Immune Deficiency, Multiple Sclerosis (MS), and Chronic Kidney Disease stage 5 were the strongest risk factors for both hospitalisation (IRR 2.9-5.4) and death (IRR 2.5-5.4) (Figure 2B, 2D and Table 2).

**Table 2.** Crude incidence rates for hospitalisation and death due to COVID-19, per 1000 person-years, stratified on Charlson Comorbidity Index (CCI), time since most recent vaccination, and age. * Investigated exposure do not contribute to CCI in stratification. CDK; Chronic Kidney Disease, compl.; complication.

|  |  |  | Hospitalisation<br>per 1000 person years |  |  |  |  |  | Death<br>per 1000 person years |  |  |  |  |  |
| --- | --- | --- | --- | --- | --- | --- | --- | --- | --- | --- | --- | --- | --- | --- |
|  |  |  | Vaccination <180<br>days |  |  | Not vaccination <180<br>days |  |  | Vaccination <180<br>days |  |  | Not vaccination<br><180 days |  |  |
|  |  |  | 18-64 | 65-79 | 80+ | 18-64 | 65-79 | 80+ | 18-64 | 65-79 | 80+ | 18-64 | 65-79 | 80+ |
| CCI* 0 | No exposure |  | 0,10 | 0,63 | 5,15 | 0,13 | 1,74 | 11,28 | 0 | 0,09 | 1,50 | 0 | 0,19 | 2,49 |
|  | Any exposure |  | 0,20 | 1,16 | 6,65 | 0,20 | 3,04 | 15,15 | 0 | 0,09 | 1,56 | 0,01 | 0,28 | 3,30 |
|  | Obesity |  | 0,81 | 2,52 | 8,61 | 0,62 | 6,80 | 13,27 | 0 | 0,34 | 1,23 | 0,01 | 1,13 | 0 |
|  | Heart Failure |  | 3,64 | 5,09 | 15,53 | 2,53 | 11,06 | 30,70 | 0 | 0,55 | 4,66 | 0,28 | 1,84 | 7,69 |
|  | Immune Deficiency |  | 4,89 | 7,65 | 16,36 | 4,63 | 22,65 | 33,97 | 0 | 0 | 0,74 | 0,99 | 1,80 | 0 |
|  | Ischemic Heart Disease |  | 0,33 | 2,19 | 11,24 | 0,56 | 6,40 | 23,94 | 0,11 | 0,05 | 2,02 | 0 | 0,42 | 4,06 |
|  | Recent Chemotherapy |  | 38,66 | 14,89 | 40,89 | 37,38 | 56,66 | 42,66 | 0 | 0 | 0 | 0 | 0 | 0 |
|  | Chronic Liver Disease |  | 1,53 | 1,18 | 3,53 | 0,60 | 10,20 | 41,30 | 0 | 0 | 0 | 0 | 1,45 | 0 |
|  | Multiple Sclerosis |  | 6,99 | 11,12 | 5,00 | 4,39 | 15,13 | 37,01 | 0 | 1,17 | 5,00 | 0 | 0 | 18,22 |
|  | Psychiatric Disorder |  | 1,35 | 4,53 | 7,05 | 0,93 | 10,21 | 19,70 | 0 | 0,59 | 5,02 | 0 | 1,27 | 9,77 |
|  | Rheumatoid Arthritis |  | 0,80 | 4,85 | 18,13 | 1,05 | 7,33 | 19,26 | 0 | 0,61 | 1,73 | 0,10 | 1,74 | 6,37 |
|  | Diabetes Mellitus | No complications | 0,47 | 2,23 | 8,65 | 0,80 | 5,78 | 19,40 | 0 | 0,16 | 1,92 | 0,07 | 0,55 | 2,24 |
|  |  | Complications | 1,09 | 3,17 | 12,29 | 1,57 | 8,26 | 29,07 | 0 | 0,08 | 2,36 | 0 | 0,77 | 3,02 |
|  | Hypertension | Medication only | 0,31 | 1,20 | 5,91 | 0,48 | 3,46 | 14,78 | 0 | 0,06 | 1,48 | 0,04 | 0,29 | 3,63 |
|  |  | Essential I10 | 1,07 | 2,48 | 9,95 | 0,94 | 6,34 | 20,49 | 0,04 | 0,17 | 1,60 | 0,02 | 0,69 | 3,96 |
|  |  | Secondary/compl | 5,38 | 0 | 14,53 | 0 | 0 | 0 | 0 | 0 | 0 | 0 | 0 | 0 |
|  | Pulmonary Disease | Mild | 0,48 | 1,78 | 7,92 | 0,53 | 5,73 | 16,53 | 0 | 0,13 | 1,58 | 0,02 | 0,26 | 3,40 |
|  |  | Severe | 3,88 | 9,36 | 18,36 | 5,33 | 22,46 | 46,60 | 0 | 0,60 | 2,08 | 0 | 2,11 | 9,16 |
|  | Chronic Kidney Disease | CKD 3 | 3,07 | 2,30 | 18,85 | 4,27 | 12,55 | 36,28 | 0 | 0 | 0 | 0 | 0 | 0 |
|  |  | CKD 4 | 0 | 8,27 | 9,97 | 0 | 17,16 | 35,91 | 0 | 0 | 2,49 | 0 | 5,68 | 8,90 |
|  |  | CKD 5 | 0 | 0 | 0 | 50,50 | 0 | 39,77 | 0 | 0 | 12,84 | 0 | 0 | 39,74 |
| CCI* 1-2 | No exposure |  | 0,95 | 3,28 | 11,95 | 0,85 | 5,25 | 23,58 | 0,12 | 0,75 | 4,36 | 0,03 | 1,24 | 7,56 |
|  | Any exposure |  | 1,90 | 4,93 | 15,36 | 1,62 | 10,72 | 28,84 | 0,08 | 0,62 | 3,75 | 0,07 | 1,45 | 6,57 |
|  | Obesity |  | 2,69 | 8,73 | 14,29 | 3,10 | 16,71 | 16,55 | 0,18 | 0,45 | 1,29 | 0,15 | 1,44 | 7,04 |
|  | Heart Failure |  | 3,75 | 10,18 | 26,32 | 5,10 | 21,57 | 48,25 | 0 | 1,44 | 5,84 | 0,38 | 3,41 | 9,52 |
|  | Immune Deficiency |  | 18,00 | 16,96 | 28,92 | 12,55 | 27,84 | 39,13 | 0,31 | 2,71 | 7,09 | 0,72 | 3,73 | 10,98 |
|  | Ischemic Heart Disease |  | 2,72 | 7,66 | 19,74 | 3,79 | 15,51 | 37,20 | 0 | 0,70 | 4,94 | 0 | 2,23 | 8,19 |
|  | Recent Chemotherapy |  | 18,07 | 44,72 | 74,15 | 26,68 | 58,11 | 81,74 | 0 | 11,03 | 27,35 | 0 | 10,38 | 53,43 |
|  | Chronic Liver Disease |  | 3,50 | 8,87 | 14,71 | 4,94 | 7,70 | 31,64 | 0 | 1,77 | 2,44 | 0 | 0 | 0 |
|  | Multiple Sclerosis |  | 14,03 | 24,11 | 17,81 | 16,10 | 36,32 | 24,35 | 0 | 4,21 | 5,90 | 0 | 0 | 24,32 |
|  | Psychiatric Disorder |  | 6,08 | 10,54 | 13,78 | 4,96 | 22,20 | 46,08 | 0,47 | 1,17 | 10,28 | 0,20 | 3,04 | 17,50 |
|  | Rheumatoid Arthritis |  | 3,38 | 9,18 | 23,97 | 2,56 | 22,76 | 38,39 | 0 | 1,33 | 5,60 | 0 | 3,95 | 7,83 |
|  | Diabetes Mellitus | No complications | 4,49 | 6,34 | 16,21 | 2,40 | 14,63 | 26,76 | 0,26 | 0,89 | 4,29 | 0,27 | 2,21 | 7,00 |
|  |  | Complications | 5,70 | 12,98 | 20,36 | 6,23 | 21,01 | 34,97 | 0 | 0,92 | 3,54 | 0 | 2,70 | 11,89 |
|  | Hypertension | Medication only | 2,50 | 3,87 | 14,17 | 2,33 | 8,53 | 27,09 | 0,16 | 0,58 | 3,23 | 0,05 | 1,17 | 7,04 |
|  |  | Essential I10 | 3,07 | 6,17 | 16,22 | 2,81 | 13,96 | 30,44 | 0,06 | 0,61 | 3,81 | 0,20 | 1,56 | 6,18 |
|  |  | Secondary/compl | 0 | 8,13 | 30,20 | 3,11 | 16,24 | 25,09 | 0 | 0 | 3,33 | 0 | 5,38 | 0 |
|  | Pulmonary Disease | Mild | 3,10 | 6,24 | 16,45 | 3,55 | 13,93 | 28,95 | 0,21 | 0,56 | 4,46 | 0,11 | 1,46 | 7,13 |
|  |  | Severe | 8,32 | 15,42 | 31,96 | 10,13 | 27,80 | 59,42 | 0 | 1,67 | 5,67 | 2,40 | 4,16 | 12,82 |
|  | Chronic Kidney Disease | CKD 3 | 5,63 | 8,78 | 17,64 | 3,13 | 27,55 | 53,46 | 0 | 0 | 5,33 | 0,78 | 1,82 | 2,63 |
|  |  | CKD 4 | 13,23 | 14,11 | 24,36 | 8,07 | 24,21 | 39,37 | 0 | 1,87 | 2,01 | 0 | 4,77 | 17,63 |
|  |  | CKD 5 | 19,98 | 18,67 | 28,17 | 18,67 | 14,30 | 58,35 | 3,06 | 0 | 6,22 | 0 | 4,73 | 11,42 |
| CCI* 3+ | No exposure |  | 2,25 | 5,41 | 16,21 | 3,92 | 11,88 | 35,15 | 0,50 | 1,62 | 3,32 | 0,10 | 0 | 6,93 |
|  | Any exposure |  | 8,30 | 14,01 | 28,33 | 7,58 | 27,45 | 47,27 | 0,29 | 1,62 | 6,43 | 0,42 | 3,10 | 12,36 |
|  | Obesity |  | 16,07 | 20,80 | 27,13 | 12,67 | 34,22 | 42,77 | 0,62 | 2,58 | 5,01 | 0,79 | 2,52 | 6,66 |
|  | Heart Failure |  | 26,45 | 26,34 | 41,06 | 17,60 | 46,22 | 54,56 | 0 | 3,80 | 9,39 | 0,51 | 6,84 | 15,40 |
|  | Immune Deficiency |  | 29,01 | 38,73 | 55,03 | 24,21 | 52,05 | 70,00 | 0 | 4,63 | 13,25 | 1,79 | 9,12 | 17,76 |
|  | Ischemic Heart Disease |  | 23,40 | 19,71 | 33,31 | 16,07 | 37,53 | 53,45 | 0,61 | 2,28 | 7,19 | 0 | 4,09 | 14,39 |
|  | Recent Chemotherapy |  | 20,03 | 18,86 | 48,27 | 22,73 | 67,92 | 73,53 | 0 | 0 | 4,78 | 0 | 0 | 28,74 |
|  | Chronic Liver Disease |  | 3,70 | 18,49 | 38,23 | 7,92 | 29,91 | 37,95 | 3,70 | 0,50 | 11,36 | 0,72 | 4,08 | 18,64 |
|  | Multiple Sclerosis |  | 16,19 | 34,88 | 51,69 | 43,33 | 103,17 | 0 | 0 | 3,85 | 0 | 0 | 37,25 | 0 |
|  | Psychiatric Disorder |  | 7,80 | 27,89 | 46,07 | 9,32 | 62,82 | 53,72 | 0 | 5,71 | 11,33 | 2,66 | 5,94 | 15,13 |
|  | Rheumatoid Arthritis |  | 5,47 | 27,24 | 49,93 | 19,49 | 53,10 | 56,64 | 0 | 3,00 | 11,02 | 0 | 12,33 | 11,98 |
|  | Diabetes Mellitus | No complications | 9,61 | 15,38 | 28,30 | 12,05 | 30,62 | 44,74 | 0,56 | 1,47 | 7,21 | 1,71 | 2,67 | 12,33 |
|  |  | Complications | 29,25 | 24,45 | 39,56 | 23,45 | 47,94 | 56,89 | 0,83 | 3,31 | 10,73 | 0,63 | 7,74 | 18,29 |
|  | Hypertension | Medication only | 10,00 | 11,09 | 28,74 | 6,21 | 25,80 | 43,60 | 0,29 | 1,33 | 6,89 | 1,13 | 2,19 | 11,31 |
|  |  | Essential I10 | 12,01 | 16,24 | 29,14 | 11,23 | 30,43 | 49,46 | 0,45 | 1,70 | 6,36 | 0,30 | 3,88 | 12,66 |
|  |  | Secondary/compl | 9,29 | 16,41 | 22,29 | 0 | 31,33 | 55,21 | 0 | 3,26 | 14,73 | 0 | 7,72 | 0 |
|  | Pulmonary Disease | Mild | 17,71 | 14,06 | 32,04 | 10,23 | 32,99 | 39,71 | 0,55 | 1,57 | 7,78 | 0,39 | 4,19 | 11,39 |
|  |  | Severe | 22,81 | 27,73 | 46,66 | 25,82 | 46,36 | 69,64 | 0 | 2,90 | 8,88 | 4,26 | 5,36 | 21,30 |
|  | Chronic Kidney Disease | CKD 3 | 13,42 | 22,41 | 45,45 | 13,68 | 39,22 | 44,71 | 0 | 1,42 | 11,22 | 1,51 | 8,71 | 10,12 |
|  |  | CKD 4 | 37,14 | 29,81 | 40,07 | 13,25 | 48,10 | 55,99 | 3,06 | 1,97 | 10,88 | 2,19 | 7,10 | 13,65 |
|  |  | CKD 5 | 36,05 | 49,32 | 53,20 | 34,37 | 68,33 | 46,58 | 0 | 7,76 | 6,44 | 0 | 14,77 | 17,46 |

Vaccination within 6 months was associated to a reduced risk of hospitalisation in multivariable regression analyses, whereas individuals with more distant (13-24 months) vaccination had a 1.9-fold [95% CI 1.8-2.0] increased risk of hospitalisation and a 2.1-fold [95% CI 1.8 - 2.5] increased risk of death. In univariate analysis (crude incidence rates), recent vaccination was associated with reduced rates of hospitalisation across the three age groups of the study (Table 2).

The present study found limited impact of socioeconomic factors for hospitalisation and death due to COVID-19 (Figure 2A, 2C).

## Discussion

The emergence of the Omicron variant of SARS-CoV-2 has significantly altered the risk landscape attributed to COVID-19. Accumulation of immunity through vaccination and natural infection, along with the reduced virulence of the Omicron strain has led to a decline in overall hospitalisation and death rates as compared to earlier phases of the pandemic.

However, certain groups remain vulnerable to severe disease and may benefit from antiviral treatment and vaccination. This study presents risk factors associated to hospitalisation and death for COVID-19 in the Swedish population after six months with Omicron circulation, mirroring the decreased risk pattern seen after the transition from pandemic to endemic SARS-CoV-2 circulation.

In the present study, increasing age was found to be the single most important predictor of both hospitalisation and death in a period with widespread population-level immunity. This finding is in line with other studies on disease severity of COVID-19 during the Omicron period (10–12).

The present study also found that advanced comorbidity played the second most critical role in explaining hospitalisation risk. This aligns with reports by Scott et al. (13), who noted that >98% of hospitalised Omicron patients in their cohort had two or more high-risk conditions. Although Omicron may exhibit lower virulence in a general sense, individuals with significant comorbidity appear to remain highly susceptible to develop severe COVID-19 also in the Omicron era. Notably, we observed recent chemotherapy, Immune Deficiency, MS, and Chronic Kidney Diseases stage 5 to be especially potent contributors to both hospitalisation and death. These findings concur with prior work from Ofer et al. (14), who showed that active cancer treatment or immunosuppressive states are consistently associated with worse COVID-19 outcomes in the Omicron period. Our data suggest that severe immunocompromising conditions and advanced organ dysfunction collectively confer an elevated vulnerability in these populations. Regarding MS, it should be noted that these patients frequently are B-cell suppressed, due to rituximab treatment that cannot be detected in our current national health registers. We hypothesise that the risk estimates associated with a MS diagnosis likely reflect the impact of rituximab treatment.

For prediction of death, care dependency emerged as the second most influential risk factor in the present study. Care dependency may reflect functional impairment and frailty, which in turn correlate with poorer resilience against infectious stressors. This observation parallels Spreco et al. (12), who reported that most fatal COVID-19 cases involved patients with very low functional levels, often dependent on nursing home care. Similarly, other studies, such as Hippisley-Cox et al. (11), have highlighted the added prognostic value of frailty indices and performance status in predicting outcome.

The present study was not designed to evaluate the effect of vaccination in specific subgroups of individuals, although our unadjusted analysis (crude incidence rates, Table 2) showed generally lower rates of hospitalisation and death in recently vaccinated individuals. Tomioka et al. (15), found that recent vaccination was particularly beneficial for individuals ≥ 80 years of age. However, several factors may negatively impact vaccine response, including immunosuppressive medication and renal failure, both of which are linked to an increased risk of hospitalisation and death due to COVID-19 in the presented material. Therefore, we suggest that additional interventions, such as early antiviral treatment should be considered for the patient groups with increased risk of severe COVID-19.

The study has some strengths. First, we have used a large sample size, improving accuracy in our estimates. Second, the use of national health registers with mandatory reporting of all cases has further improved our accuracy and minimised selection biases. Furthermore, high-coverage registers and investigation of the entire population ensures high external validity.

The study has several limitations. First, hospitalisation due to COVID-19 does not always indicate severe illness, as very frail individuals may require hospital admission even for mild disease. Additionally, a potential bias may have been introduced in the hospitalisation analysis, as patients with immunosuppressive conditions are more likely to be admitted for Remdesivir treatment, and hospital admission is standard practice for patients on chemotherapy that present with fever. However, this bias would not affect death analyses, which generally showed patterns similar to those of the hospitalisation analyses. Second, the use of individual models for each assessed comorbidity to better assess confounding, may have introduce some variation in the underlying risk of the reference group. Third, we do not have access to data on immunosuppressive treatments given at hospital apart from chemotherapy, for example to patients with MS and Rheumatoid Arthritis. However, regardless of the underlying mechanisms, the presented estimates accurately reflect the real-world situation patients with the investigated diagnoses. Forth, as there was no public testing for SARS-CoV-2 during the study period, COVID-19 deaths outside of hospitals may have been underreported.

In conclusion, although the Omicron variant generally exhibits lower virulence than previous SARS-CoV-2 variants, vulnerable individuals remain at elevated risk of developing severe COVID-19. Our study showed that advanced age is still the strongest risk factor for hospitalisation and death due to COVID-19, followed by advanced comorbidity and care dependency. These findings can guide recommendations for vaccination strategies and antiviral treatment, helping to protect those most at risk.

## Data availability

The data underlying this article cannot be shared publicly due to regulations under Swedish law. According to the Swedish Ethics Review Act, the General Data Protection Regulation, and the Public Access to Information and Secrecy Act, patient data can only be made available, after legal review, to researchers who meet the criteria for access to this type of confidential data. Requests regarding data in this report may be made to the corresponding author.

---

**Supplementary Table S1.**
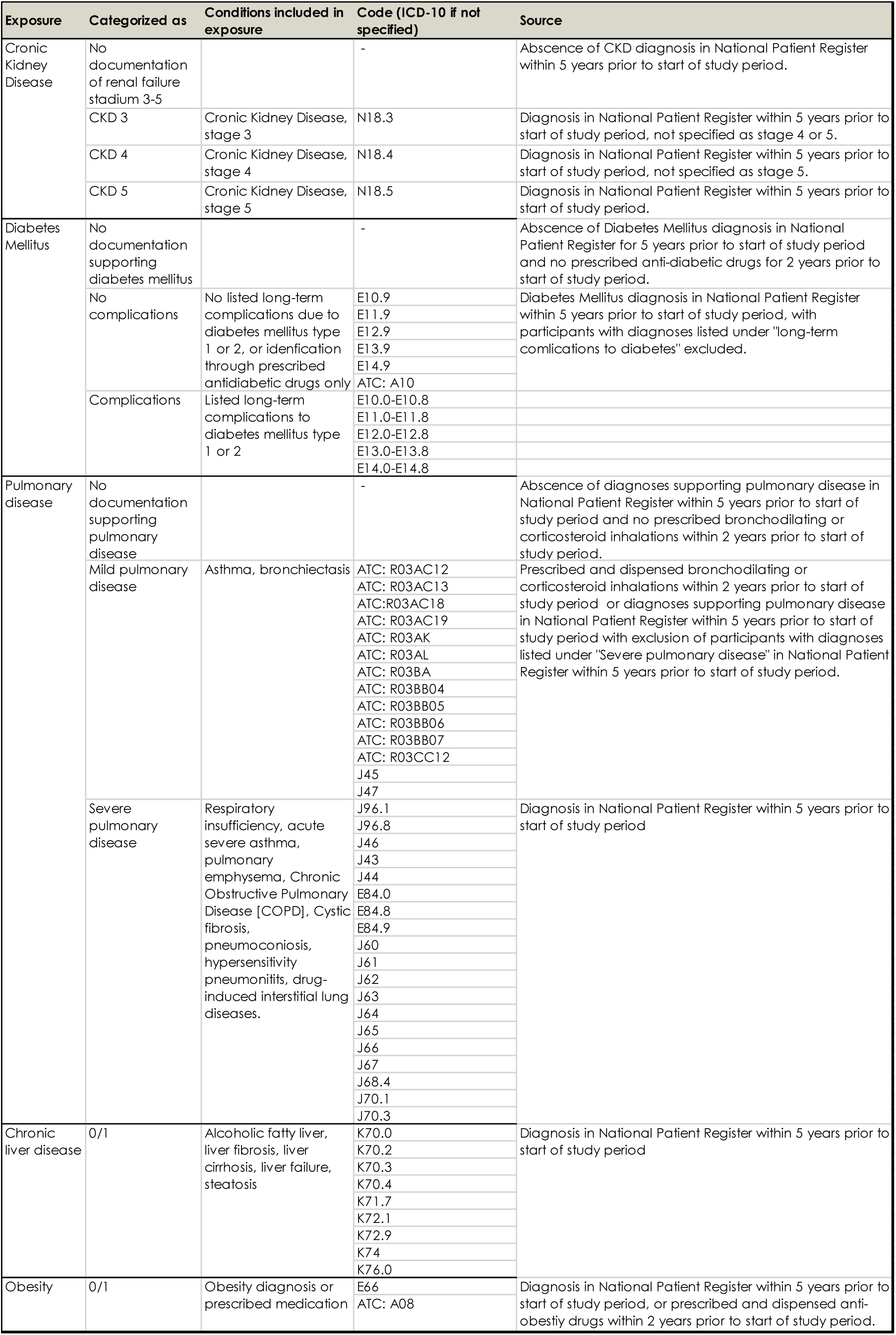
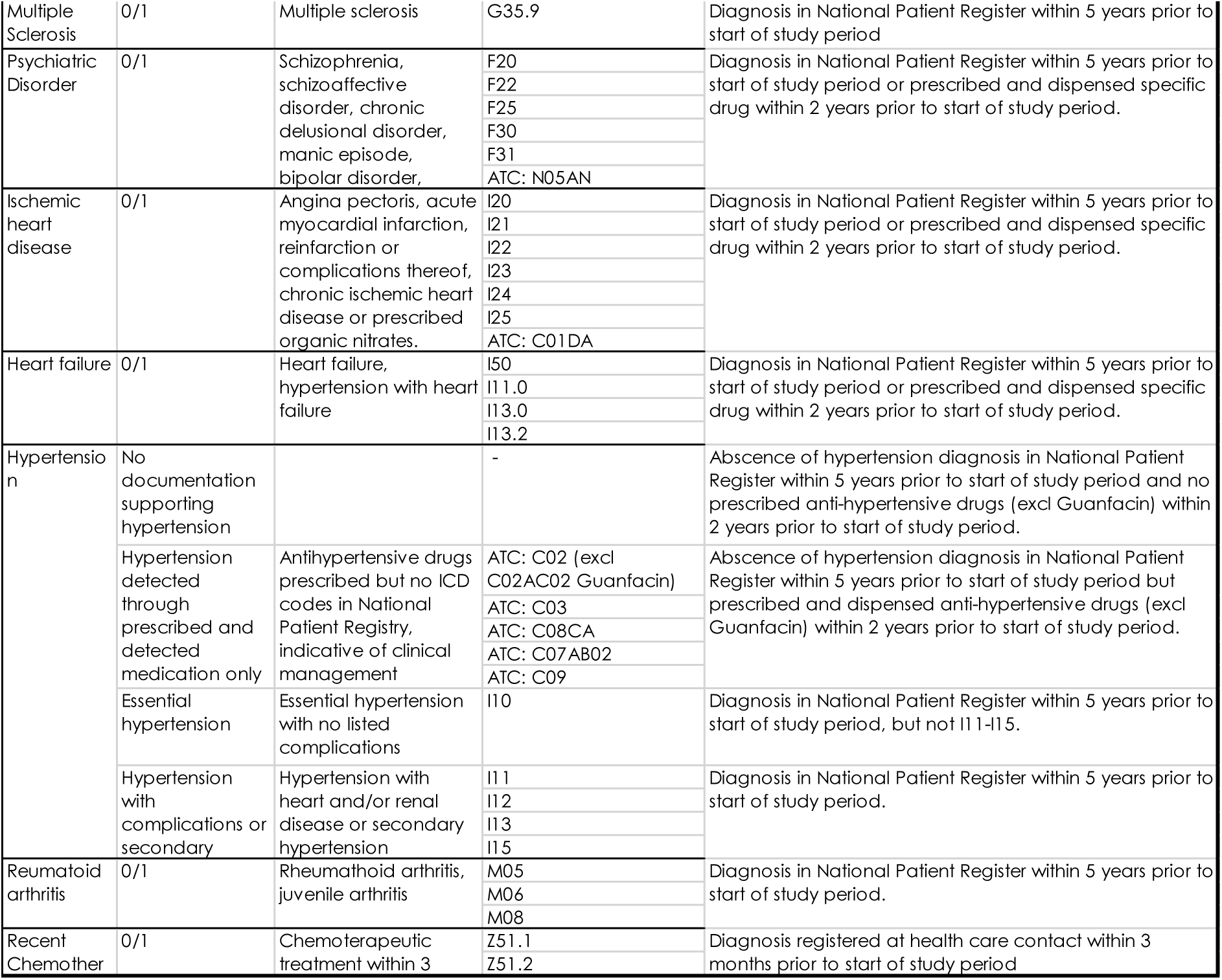
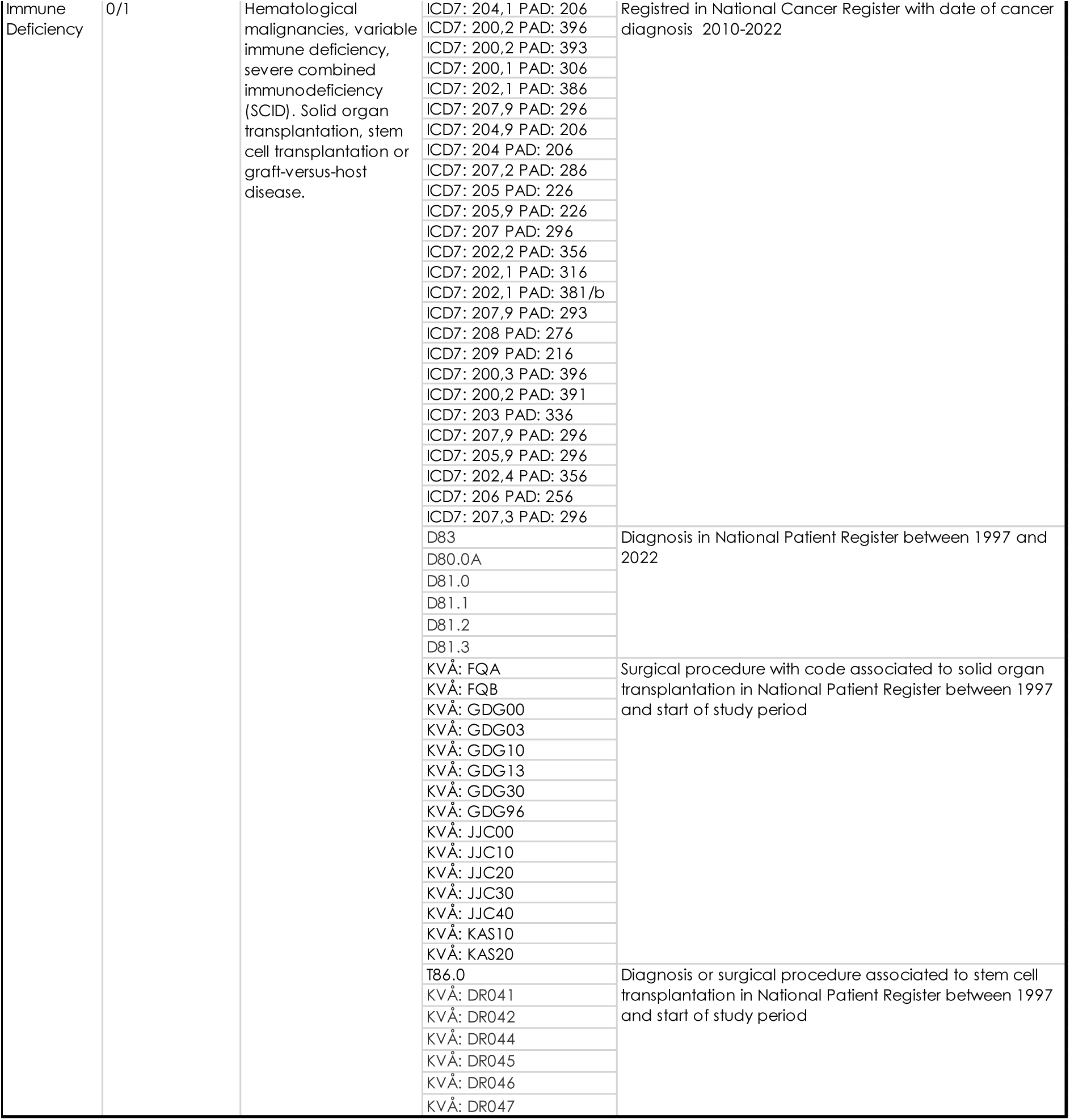
Categorisation of health conditions identified through codes of the 10^th^ International Statistical Classification of Diseases (ICD-10) classification, Anatomic Therapeutic Chemical (ATC) classification, and health care procedures (“KVÅ” codes), in national health registers.

**Table S2.** STROBE Statement—Checklist of items that should be included in reports of *cohort studies*.

|  | Item No | Recommendation | Comment |
| --- | --- | --- | --- |
| Title and abstract | 1 | (a) Indicate the study’s design with a commonly used term in the title or the abstract | Title |
|  |  | (b) Provide in the abstract an informative and balanced summary of what was done and what was found | Abstract |
| Introduction |  |  |  |
| Background/rationale | 2 | Explain the scientific background and rationale for the investigation being reported | Introduction |
| Objectives | 3 | State specific objectives, including any prespecified hypotheses | Introduction |
| Methods |  |  |  |
| Study design | 4 | Present key elements of study design early in the paper | Abstract and Methods |
| Setting | 5 | Describe the setting, locations, and relevant dates, including periods of recruitment, exposure, follow-up, and data collection | Methods |
| Participants | 6 | (a) Give the eligibility criteria, and the sources and methods of selection of participants. Describe methods of follow-up | Methods |
|  |  | (b) For matched studies, give matching criteria and number of exposed and unexposed | - |
| Variables | 7 | Clearly define all outcomes, exposures, predictors, potential confounders, and effect modifiers. Give diagnostic criteria, if applicable | Methods and Suppl material |
| Data sources/measurement | 8* | For each variable of interest, give sources of data and details of methods of assessment (measurement). Describe comparability of assessment methods if there is more than one group | Methods and suppl material |
| Bias | 9 | Describe any efforts to address potential sources of bias | Methods |
| Study size | 10 | Explain how the study size was arrived at | Methods |
| Quantitative variables | 11 | Explain how quantitative variables were handled in the analyses. If applicable, describe which groupings were chosen and why | Methods |
| Statistical methods | 12 | (a) Describe all statistical methods, including those used to control for confounding | Methods |
|  |  | (b) Describe any methods used to examine subgroups and interactions | Methods |
|  |  | (c) Explain how missing data were addressed | Methods |
|  |  | (d) If applicable, explain how loss to follow-up was addressed | Not applicable |
|  |  | (e) Describe any sensitivity analyses | Methods |
| **Results** |  |  |  |
| Participants | 13* | (a) Report numbers of individuals at each stage of study—eg numbers potentially eligible, examined for eligibility, confirmed eligible, included in the study, completing follow-up, and analysed | Results |
|  |  | (b) Give reasons for non-participation at each stage | Not applicable |
|  |  | (c) Consider use of a flow diagram | Not applicable |
| Descriptive data | 14* | (a) Give characteristics of study participants (eg demographic, clinical, social) and information on exposures and potential confounders | Results, table 1 |
|  |  | (b) Indicate number of participants with missing data for each variable of interest | Results |
|  |  | (c) Summarise follow-up time (eg, average and total amount) | Results |
| Outcome data | 15* | Report numbers of outcome events or summary measures over time | Results |
| Main results | 16 | (a) Give unadjusted estimates and, if applicable, confounder-adjusted estimates and their precision (eg, 95% confidence interval). Make clear which confounders were adjusted for and why they were included | Results |
|  |  | (b) Report category boundaries when continuous variables were categorized | Results and Methods |
|  |  | (c) If relevant, consider translating estimates of relative risk into absolute risk for a meaningful time period | Results |
| Other analyses | 17 | Report other analyses done—eg analyses of subgroups and interactions, and sensitivity analyses | Results |
| **Discussion** |  |  |  |
| Key results | 18 | Summarise key results with reference to study objectives | Discussion |
| Limitations | 19 | Discuss limitations of the study, taking into account sources of potential bias or imprecision. Discuss both direction and magnitude of any potential bias | Discussion |
| Interpretation | 20 | Give a cautious overall interpretation of results considering objectives, limitations, multiplicity of analyses, results from similar studies, and other relevant evidence | Discussion, conclusion |
| Generalisability | 21 | Discuss the generalisability (external validity) of the study results | Discussion |
| **Other information** |  |  |  |
| Funding | 22 | Give the source of funding and the role of the funders for the present study and, if applicable, for the original study on which the present article is based | Funding |

\*Give information separately for exposed and unexposed groups.
**Note:** An Explanation and Elaboration article discusses each checklist item and gives methodological background and published examples of transparent reporting. The STROBE checklist is best used in conjunction with this article (freely available on the Web sites of PLoS Medicine at http://www.plosmedicine.org/, Annals of Internal Medicine at http://www.annals.org/, and Epidemiology at http://www.epidem.com/). Information on the STROBE Initiative is available at http://www.strobe-statement.org.

